# The new SARS-CoV-2 variant and reinfection in the resurgence of COVID-19 outbreaks in Manaus, Brazil

**DOI:** 10.1101/2021.03.25.21254281

**Authors:** Daihai He, Guihong Fan, Xueying Wang, Yingke Li, Zhihang Peng

## Abstract

Manaus, a city of 2.2 million population, the capital of Amazonas state of Brazil was hit badly by two waves of COVID-19 with more than 10,000 severe acute respiratory syndrome deaths by the end of February 2021. It was estimated that the first wave infected over three quarters of the population in Manaus based on routine blood donor data, and the second wave was largely due to reinfection with a new variant named P1 strain. In this work, we revisit these claims, and discuss biological constraints. In particular, we model the two waves with a two-strain model without a significant proportion of reinfections.

## Introduction

Antibody sero-prevalence is one commonly used approach in detecting infection attack rate (IAR) since infected people would normally show a positive antibody in their blood if they have ever being infected no matter symptomatic or asymptomatic infection (*1*). A study on COVID-19 sero-prevalence (proportion of sero-positive) among routine blood donors in two Brazilian cities: Manaus and Sao Paulo caught wide media attention (*2*). The study reported a high IAR of COVID-19 among routine blood donors, around 75% by October 2020, in Manaus, a city of 2.2 million population in Amazonas, Brazil. It was further generalized to the whole population of the city and concluded that the IAR exceeded 75% by October 2020, which left the city with at most 25% of people uninfected after the first wave of the pandemic. Unexpectedly, the city was hit by an even larger second wave between December 2020 and February 2020. The team (*2*) argued that the second wave was largely caused by reinfection (mainly of a new strain, named P1 strain), since given 75% IAR in the first wave and an even larger second wave in Manaus (with a hypothetical IAR >75% for the second wave), there must be approximately 50% of the population that were infected twice, once in the first wave and again in the second wave. This means around 1 million people were infected twice. With such a big re-infected population, the chance will be high to find a large number of reinfections. However, according to official reports (*3*) there were only 3 reinfections confirmed in the state of Amazonas (4 million population including the capital Manaus). In other regions, reinfection is generally rare (*4, 5*). If a substantial amount of reinfections cannot be found in Manaus, then the blood donor study (*2*) most likely overestimated the population sero-prevalence. Indeed, a population sero-prevalence study in 133 Brazilian cities including Manaus (*6*) found a much smaller (∼20%) sero-prevalence in May-June, 2020. Hence, the blood donor data in Manaus (*2*) are most likely biased and/or their analyses are flawed (*7*). In this work. we revisit the blood donor data in Manaus with the aim to reveal the possible reasons behind the discrepancy.

We first obtain the observed sero-prevalence among blood donors from (*2*), with 95% confidence intervals, after an adjustment for age-sex of the sample, and sensitivity and specificity of tests. We display the observed sero-prevalence with dashed lines in Figure 1a (those from Manaus in red, while those from Sao Paulo in black). The observed sero-prevalence showed a wave pattern in Manaus. To explain this wave pattern, Buss et al. proposed a sero-reversion mechanism, namely, the antibody level of a recovered sero-positive person may decay and turn sero-negative over time. Hypothetically, if there was no waning of antibody and/or sero-reversion, the population sero-prevalence should be a non-decreasing function of time. The wave pattern of sero-prevalence is a result of sero-conversion (due to new infection) and sero-reversion (due to waning of antibody). Since the sero-conversion part is a result of new infections, it should be connected to reported cases or deaths. However, Buss et al. did not incorporate the whole time series of reported COVID-19 cases and deaths when they estimated the sero-reversion function, but only used the fact that the reported cases and deaths were low during July and August 2020. The reason behind exclusion of the whole time series could be due to the quality of the reported cases and deaths (low reporting rate). Then it is unclear how well their results on sero-prevalence match the epidemic embedded in the reported cases and deaths. To resolve this issue, we incorporate the reported Severe Acute Respiratory Syndrome (SARI) deaths which is largely attributed to COVID-19 infection (from observation in all regions, COVID-19 is virtually the only respiratory infections during the height of the pandemic). Using the SARI deaths, instead of reported COVID-19 deaths, resolves the issue of under reporting to some extent. With blood donor sero-prevalence data and SARI deaths, we aim to explore how the explanation of the observed sero-prevalence in the blood donors is dependent on biological assumptions. We also show the possibilities to explain these blood donors data, the two waves of deaths and the emergence of the new variant without a significant proportion of reinfections.

**Figure 1:**
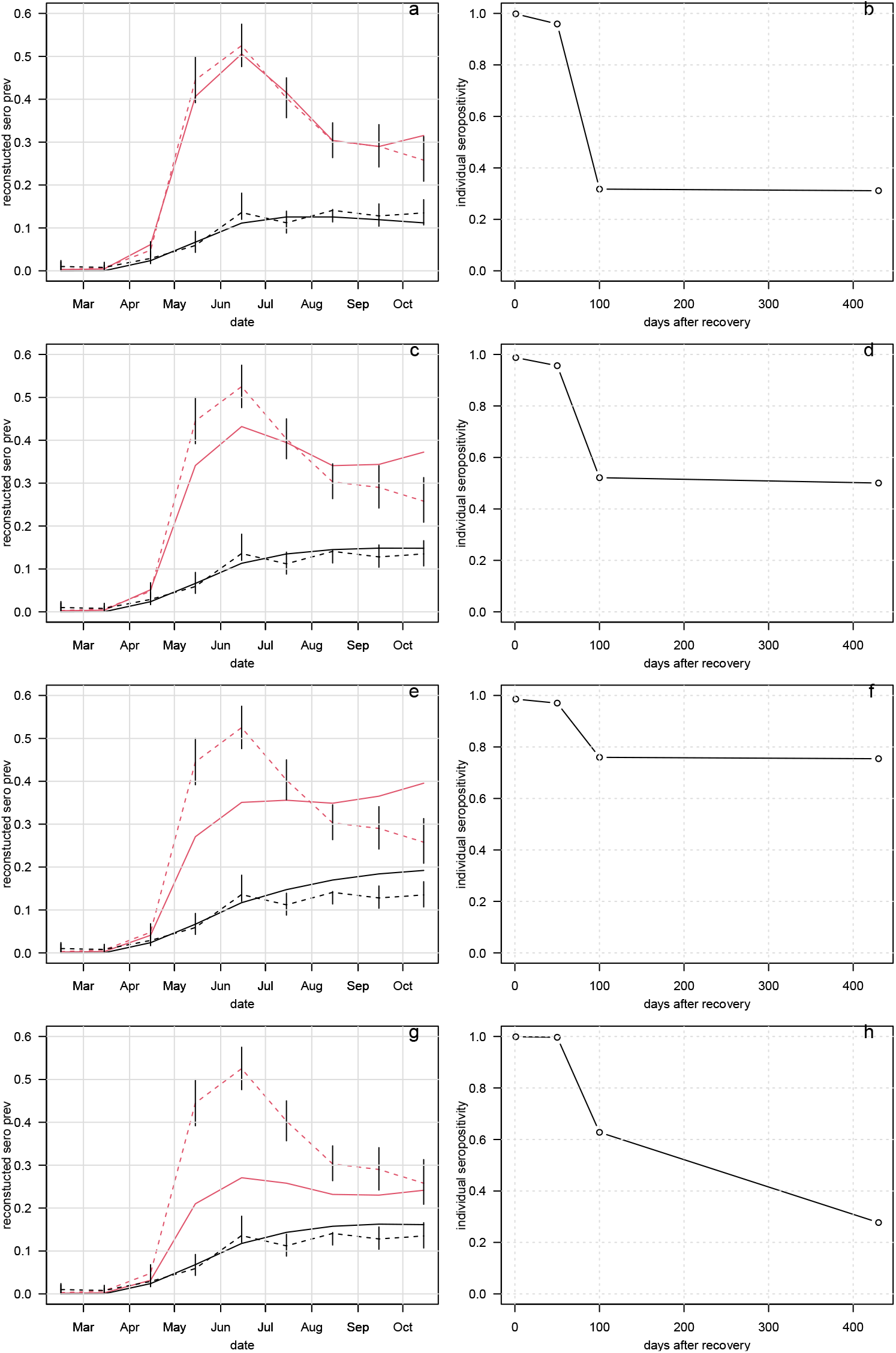
Reconstruct blood donor sero-prevalence. Left panels compare the observed (dashed) and reconstructed (solid) sero-prevalence of blood donors in Manaus (red curves) and San Paulo (black curves). Right panels show the four scenarios of reconstructed antibody decay functions of individual sero-positivity since recovery. Panel (g,h) show the case of k<1.5. To achieve a reasonable fit, one needs a rapid drop of seropositivity and large k=2.94 in (a,b).

## Methods

Denote the daily COVID-19 antibody sero-prevalence (those defined (*2*)) as *p*, daily sero-conversion as *p*^*+*^, daily sero-reversion as *p*^*-*^, then we have the following equation:

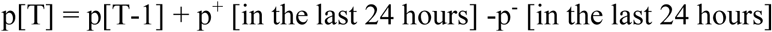

This can be written as

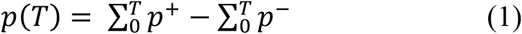

Denote f(s) as the decay function of individual seropositive with 0<f(s)<1, which is a monotonically decreasing function of time s (time since the recovery). Denote d(t) as the time series of SARI deaths (a good proxy of true COVID-19 deaths). Denoted r as the infection fatality ratio (IFR). The sero-prevalence on day T is given by the following the following convolution (here we rewrite the summation in the above equation into integral fashion) between d(t) and f(s):

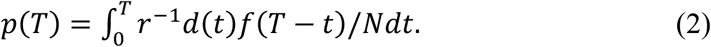

Note that here we assume a 100% sero-conversion ratio (per infection) for the sake of simplicity, but in reality this ratio is less than 1 but close to 1. N is the size of population. We assume a constant IFR between daily SARI deaths and COVID-19 infections. One death reported in a day implies 1/IFR (or 1/IFR-1) sero-conversions of infected and recovered cases, and 1/IFR≫ 1. Here we assume the delay between infection and death (∼14 days) equals the delay between infection and sero-conversion for recovered (∼14 days). We assume a piece-wise linear function for f(s). The function is determined by probabilities of sero-positivity at day 0, 50, 100 and 400 for a typically recovered individual, denoted as p_1_, p_2_, p_3_, and p_4_, respectively. We assume a decreasing trend p_1_>p_2_>p_3_>p_4_, and estimate p_1_ to p_4_ with some restrictions on residual positivity p_4_>0.25, p_4_>0.5 or p_4_>0.75. A recent study showed a significant residual positivity lasts for at least half year (*8*).

For the sake of simplicity, we assume a constant IFR= 1/120 (approximately 0.83%) for Sao Paulo which is in line with (*2, 6*). We assume Manaus has a smaller IFR (given a relatively younger population(*2*)) which is 1/k of IFR in Manaus, namely the IFR in Manaus is 0.83%/k.

We test two cases: first, k is to be estimated freely, second, k<1.5.

We randomly sample parameters (k, p_1_ to p_4_) for a large number of (e.g. 100,000) times, and calculate the sum of the Scaled Squared Errors (SSE, which equals the difference divided by the width of confidence interval, then squared) between the observed and the reconstructed sero-prevalence, and then minimize the SSE.

The reconstructed sero-prevalence is a convolution of the time series of daily deaths and the decay function of sero-positivity. The reconstructed (solid curve) and observed (dashed curved) are compared in Figure 1.

## Results

In Figure 1, we compare the observed sero-prevalence in blood donor and reconstructed population level sero-prevalence (using the above method), with dashed lines for observed and solid lines for reconstructed in left panels (those from Manaus in red, while those from Sao Paulo in black). Left panels show the ‘best’ fitting performances with different decay functions of individual seropositivity over time in the right panels. Residual positivity p_4_>0.25, p_4_>0.5 or p_4_>0.75 are used in panels (a,b), (c,d), (e,f), with k free, respectively. While k<1.5 with p_4_>0.25 are used in panels (g,h).

Our estimated parameter values are listed in the following table.

## Discussion and Conclusion

The first scenario (Fig. 1 a,b) yields the smallest SSE, and the best matching between observed and reconstructed sero-prevalence from SARI death data with a decay function in panel b. However, one needs to consider biological constraints. The question is whether the rapid drop of individual seropositivity is biologically reasonable and whether the significant difference in IFRs (k=2.94) between the two cities (San Paulo and Manaus) is biologically reasonable.

In the second scenario, we lift the residual seropositivity (p_4_) slightly, the fitting in Manaus worsen evidently (see Fig. 1 c,d). This indicates that the matching in (a,b) sensitively depends on this biological assumption.

In the third scenario, The fitting in Manaus completely fails in (e,f), while the fitting in Sao Paulo could be acceptable, if we assume that the blood donation screening led to a lower sero-prevalence among blood donors than in the general population (reconstructed based on death data, IFR and sero-reversion). The biological constraints on p_4_ more severely impact the data interpretation in Manaus but less so in San Paulo. In the fourth scenario, we assume the difference in IFR between the two cities are relatively small, k<1.5, and the fitting in Manaus fails. This means that the matching between observed and reconstructed in Manaus is also dependent on the assumption of a very different IFR. Although Manaus has a much younger population, we note that the fact of a much younger population could imply a poor health condition (short life expectancy), high prevalence of chronic illnesses, thus a high death rate in COVID-19 pandemic. We cannot only focus the young population side and overlook the reason behind the young population, for instance, the extreme shortage of medical supplies in Manaus during the pandemic. Young population does not automatically mean a small IFR. The fitting in Manaus sensitively depends on two biological assumptions, a rapid drop of sero-positivity and a huge difference in IFR as compared to Sao Paulo. Overall, the fitting in Sao Paulo is less sensitive to the assumption on sero-positivity.

Alternatively, there is a possibility that the blood donor data in Manaus were probably impacted severely by the peak of the pandemic (more than 100 deaths per day in a city of 2.2 million population), due to reasons such as the shortage of blood donations. Hence, some high risk group of peoples (with a high seropositivity) were recruited in blood donation and caused a higher sero-prevalence among blood donors in Manaus than that in Sao Paulo which is less severely impacted by the pandemic. It is worth pointing out that the blood donation in Brazil (especially in Amazonas) were different from other high income countries/regions.

Amazonas region has a higher family replacement donors (and a higher proportion of repeated donors). In Brazil, around 60% of their blood donors are voluntary, and other 40% are family replacement donors which means if a family member used blood storage in the blood bank, his/her relative needs to donate blood into the blood bank as a replacement. The risk of a family member of a patient might be higher than the general population. This might cause a high sero-prevalence among blood donors. Sudden high prevalence of seropositivity of other infections among donations occurred in the past in Brazil, e.g., 17.23% HBs Ag positivity in Amazonas and 18.34% syphilis positivity in Rio de Janeiro in 2014 according to 4º Boletim de Produção Hemoterápica – HEMOPROD 2014-2015 report. In view of these, the calculated 75% infection attack rate from blood donors should be cautiously generalized to the whole population. In the following, we show a hypothetical scenario which could cause the observed pattern.

### Hypothetical scenario to explain the blood donor sero-prevalence

We show that a fluctuation in the composition of blood donors (high risk versus low risk) can explain the observed fluctuation in the blood donor COVID-19 seropositivity reported in Manaus. We assume that blood donors can be divided into high risk group (socially active) and low risk group (socially not active) of roughly equal proportion. In Table 2, the first column shows month, the second column shows raw seropositivity from blood donor data. The third column shows the percentage of high risk group which could have varied during peak of pandemic, due to a shortage of spontaneous voluntary donation which was reported elsewhere, too. High risk groups, e.g. certain professions, socially active groups, people living in crowded conditions, military members, health care works, family replacement donors (*9*), could have a relatively high risk of seropositivity for infections (for family replacement donors, see(*9*)). Although there were studies which show the safety of family replacement donation in Brazil against HIV/HBV, it is unclear for COVID-19 which spreads more rapidly with a large proportion of asymptomatic or mild symptomatic cases. A deeper exam of the details of the donors composition reported in (*2*) are needed, given the importance of the topic. The fourth column shows the relative risk for COVID-19 seropositivity, where we assume a fluctuation as well to reflect the increased risk of COVID-19 infection during the peak of the pandemic. The fifth column shows the hypothetical true population sero-prevalence. The tail of the first wave did not die out during the period, but persisted at a low level, which contributed to sero-conversion on the one hand. One the other there were sero-reversion (although the rate of the decay is unknown). Considering both effects, the true population seroprevalence might be stable or slightly dropped, as we hypothesized in the first column.

**Table 1.** Estimated parameter values.

| Scenario | $p_1$ | $p_2$ | $p_3$ | $p_4$ | $k$ | SSE |
| --- | --- | --- | --- | --- | --- | --- |
| 1 | 0.9985 | 0.9591 | 0.3175 | 0.3115 | 2.9463 | 2.6108 |
| 2 | 0.9878 | 0.9563 | 0.5211 | 0.5006 | 2.4872 | 5.4082 |
| 3 | 0.9859 | 0.9704 | 0.7597 | 0.7544 | 1.9627 | 13.605 |
| 4 | 0.999 | 0.9969 | 0.6277 | 0.2772 | 1.4969 | 17.9086 |

**Table 2.** A hypothetical scenario to explain the peak in seroprevalence among blood donor.

|  | Observed | Hypothetical |
| --- | --- | --- |

| month | Raw sero-prevalence among blood donors from (2) | Proportion of high risk donors among all blood donors in Manaus, r | Relative risk ratio of high risk donor vs low risk donor, s | Hypothetical true population sero-prevalence, p |
| --- | --- | --- | --- | --- |
| 2 | 0.1 | 0.5 | 5.6 | 0.1 |
| 3 | 0.7 | 0.5 | 5.6 | 0.7 |
| 4 | 5.5 | 0.7 | 7.84 | 5 |
| 5 | 39.9 | 0.7 | 7.84 | 20 |
| 6 | 46.3 | 0.7 | 7.84 | 22 |
| 7 | 36.5 | 0.6 | 6.72 | 24 |
| 8 | 27.5 | 0.5 | 5.6 | 26 |
| 9 | 24.7 | 0.5 | 5.6 | 25 |
| 10 | 20.7 | 0.5 | 5.6 | 24 |

The blood donor screening can only exclude, to some extent, active or recent symptomatic case (and close contracts of such cases) in previous 30 days, while the sero-conversion takes weeks to peak (namely sero-positivity peaks weeks after the infection). The effectiveness of this type of screening is inefficient to exclude sero-positive cases of COVID-19. (Note that the HIV/HBV prevalence among blood donors in Brazil was two order magnitude higher than other high-income countries/regions, thus about 1/5 of blood donations are excluded due to HIV/HBV positivity and other infections.) Nevertheless, we assume the blood donors have a relative low prevalence than the true population baseline due to the screening, e.g. 1/3 of the risk of the true baseline. Low risk donor proportion equals 1 minus proportion of high risk donors. Putting all of the above together, we can simulate the observed raw sero-prevalence.

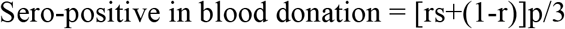

Figure 2 compares the mocked and the observed sero-prevalence among blood donors, where we achieve a reasonable matching.

**Figure 2.**
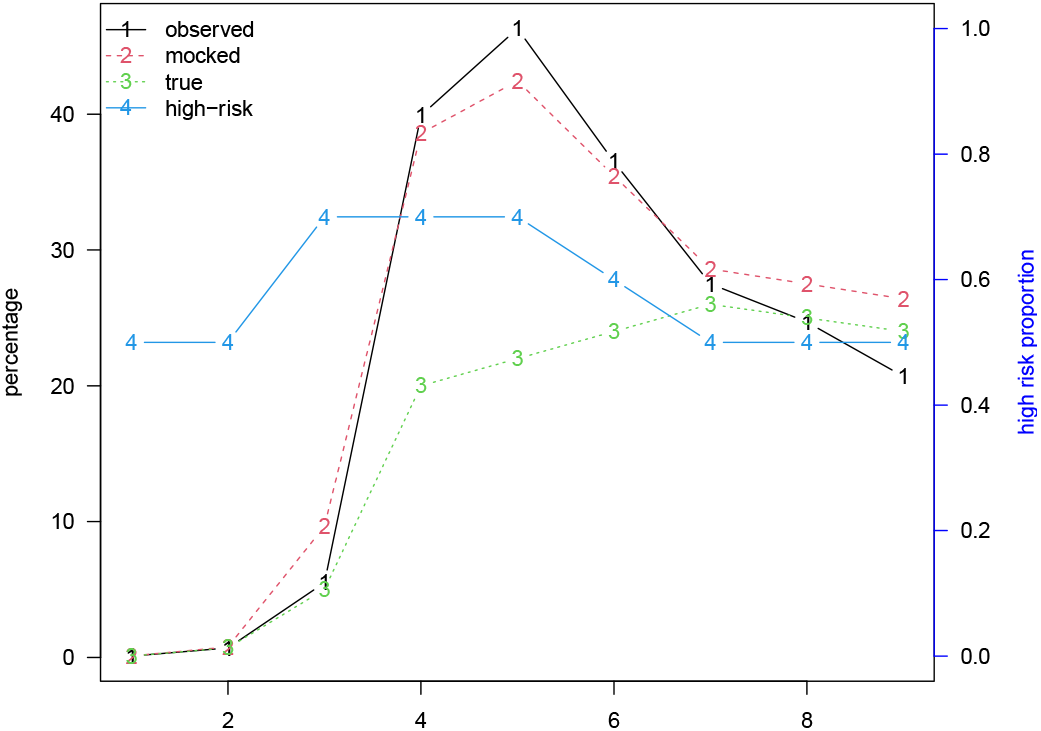
The observed raw seroprevalence and mocked seroprevalence among blood donors in Manaus, and the hypothetical true prevalence and proportion of high risk donors. We reproduce the observed sero-positivity in the blood donors with our hypothetical scenario.

### Decay rate of sero-prevalence

In a preprint, Álvarez-Antonio et al. (*10*) conducted a population-level serological study in Iquitos, Loreto, Peru in July-August, 2020. The city of a population of 460,000 was hit badly in May, 2020 with more than 2500 (confirmed and suspected) deaths. Finger-prink blood samples from a group of individuals were taken in July, then in August, 2020, with raw seroprevalence levels at 70% (July) versus 66% (August). The slight drop in the seroprevalence is likely due to seroreversion, deducted a small proportion of sero-conversion (negative in July and positive in August due to delay of sero-conversion or new infection). Nevertheless, a drop of 5.7% with respect to the baseline level in July. However, the sero-prevalence level among blood donors (*2*) peaked at May to July in Manaus and dropped at surprisingly high rate, 24.8% from July to August, which is 4.35-fold of that of the Iquitos study given similar waves. We note that different assays could behave differently, the difference is still higher than reported elsewhere(*11, 12*). Also, the seropositivity decay function requires one’s antibody to drop to low level rapidly (*2*). Further validity of this assumption is needed. A recent study (*8*) found that the level of neutralizing antibodies remained stable at follow-up periods of half of year (44.6 and 41.2 percent, respectively) in Wuhan, China.

### A two-strain model

It was reported that P1 strain could have caused the second wave in Manaus with a large amount of reinfections (*13-15*). Laboratory experiments hint the possibility of reinfection, but very few reinfections were reported so far. This discrepancy needs serious examination. We consider a scenario without reinfections given the fact that very few reinfections was reported. The reinfection is a fact, however, the contribution of reinfection needs serious evaluation.

We develop a two-strain model by adding a loop of P1 strain caused Exposed, Infectious, Hospitalized, Deaths or Recovered cases, besides a loop of non-P1 caused Exposed, Infectious, Hospitalized, Deaths or Recovered cases. P1 strain and non-P1 strain compete on the same susceptible pool. The one-strain model and our methodology was explained in a preprint(*16*). We show our fitting results in Figure 4 of (*16*), where we fitted our two-strain model to the reported SARI deaths. We assume that the transmission rate of P1 strain is 2.2-fold of that of non-P1 strain (*13*). The P1 strain was introduced in the middle of November, 2020.

Our model fitting is at least as good as those in (*13-15*) without any reinfection. Then it is questionable why we should trust a high reinfection rate estimation based similar model fitting. We argue that this kind of model fitting to SARI deaths and sequence proportion data cannot be used to detect a very weak signal of reinfection if it is indeed as low as 1% or 0.1%. If the true proportion is higher than 5% or 25%, then it is strange that only three official confirmed reinfections were reported so far. It is fine to raise the alarm of reinfection. A large-scale household study or patient record study is more powerful to determine the true rate of reinfection.

In this work, we revisit the blood donor seroprevalence study and reinfection estimation study in (*2, 14, 15*). Our conclusion is that when we infer population seroprevalence using convenient blood donor seroprevalence data we need to exam biological constraints carefully. The role of reinfection in the resurgence of COVID-19 in Manaus needs more data from large-scale household or patient record study to quantify.

## Data Availability

All data used in this work were publicly available.

## Declarations

### Ethics approval and consent to participate

Neither ethical approval nor individual consent was not applicable.

### Availability of materials

All data used in this work were publicly available.

### Consent for publication

Not applicable.

## Acknowledgements

None.

## Disclaimer

The funding agencies had no role in the design and conduct of the study; collection, management, analysis, and interpretation of the data; preparation, review, or approval of the manuscript; or decision to submit the manuscript for publication.

## Author’s contributions

DH conceived the study, and carried out the analysis. All authors discussed the results, drafted, read and revised the manuscript, and approved for publishing.

